# FERMAT: An Autoregressive Clinical Foundation Model for Disease Risk and Clinical Event Prediction

**DOI:** 10.64898/2026.10.11.26365210

**Authors:** Jaegyun Jung, Hyung-Chul Lee

## Abstract

Clinical foundation models learn patterns in longitudinal health records that can be reused across prediction tasks. However, the relative strengths of representation-based and generation-based prediction across clinical tasks remain unclear. Using longitudinal electronic health records from 3.77 million patients at a single hospital, we developed FERMAT (Foundation model for Exploring Real-world Multimodal health data using Autoregressive Trajectory modeling), a 65-million-parameter clinical foundation model. We compared predictions from task-specific predictors trained on patient representations with those derived from generated clinical event sequences. Representation-based prediction performed better in the disease-risk comparisons. In detailed analyses of five-year chronic kidney disease, fatty liver, and diabetes risk in a test cohort of 10,000 patients, representation-based predictors exceeded generation-based estimates by 0.098–0.115 in observation-weighted AUROC and had lower probability error. Generation-based prediction, in turn, performed better in predicting diagnoses, prescriptions, and procedures recorded on the same day, a task motivated by the frequent presence of multiple events with the same date in hospital EHRs. Using 50 generated trajectories per patient, it achieved a Recall@10 of 71.0%, compared with 55.7% for representation-based prediction, with a paired improvement of 15.31 percentage points (95% CI, 13.92–16.79; *n* = 1,862). These findings demonstrate task-dependent strengths of representation-based and generation-based applications within a single clinical foundation model, providing an empirical basis for exploring how the two approaches can be used individually or combined to address clinical prediction needs.

## 1 Introduction

Foundation models are pretrained on large, diverse datasets and can be adapted to a range of downstream tasks [1]. This capacity to reuse a model across tasks offers a basis for developing medical AI that supports multiple clinical needs. Clinical prediction using electronic health records (EHRs) is one potential application of foundation models in medicine. EHRs contain diagnoses, prescriptions, procedures, and other observations collected over time, providing patient histories that can be used for pretraining [2–7].

Existing EHR studies illustrate different ways to use information learned during pretraining. CLMBR and MOTOR use patient representations to train predictors for clinical outcomes and time-to-event tasks, respectively [2, 3]. In this representation-based approach, a numerical summary of the observed history serves as input to a task-specific predictor. Autoregressive models can also extend the observed history by generating clinical event sequences, from which predictions can be derived. This generation-based approach has been investigated through disease histories in Delphi-2M, medical-event sequences in Curiosity, zero-shot outcome prediction with ETHOS, and medical-concept forecasting with Foresight [4–7].

Existing comparisons have examined these applications for clinical outcomes such as disease onset and readmission [10]. Waxler et al. also evaluated predictions of diagnoses, medications, laboratory tests, and procedures recorded during a clinical encounter, comparing generated items with baselines that reuse previously recorded items [5]. However, these findings do not yet clarify how the predictive strengths of these applications vary with the clinical task and the characteristics of the underlying EHR data. For example, hospital EHRs contain multiple clinical items recorded on the same date, a feature reflected in models that represent patient histories as sequences of daily event sets [11]. Comparisons should therefore examine each application’s strengths across clinical tasks and interpret the findings in the context of such record characteristics.

Here we present FERMAT, a 65-million-parameter autoregressive clinical foundation model. We evaluate its representation-based and generation-based applications using tasks that reflect clinical prediction needs and the organization of hospital EHRs: disease-risk estimation and prediction of diagnoses, prescriptions, and procedures recorded together. By comparing the applications on the same patients, observed histories, and targets within each task, we examine their respective predictive strengths across these settings.

The two tasks ask different clinical questions. Disease-risk estimation asks for the probability of a prespecified outcome over several years, the usual target of clinical risk models. The record task asks which diagnoses, prescriptions, and procedures will be recorded together on the first clinical date after the observed history. Anticipating the content of the next encounter has been a target of EHR prediction since Doctor AI [8]; a ranked list of likely items could support preparation for that encounter, such as review of anticipated prescriptions or tests, although we did not evaluate such use. The record task also tests the first step of trajectory generation, on which longer simulations depend.

FERMAT builds on established components. Like Delphi-2M, it replaces positional encoding with continuous age, models the waiting time to the next event, and masks attention to events that share the predicted event’s time [4]. ETHOS and Curiosity instead insert discrete time-interval tokens between events and omit them for events closer together than the smallest interval, so whether time passes is decided implicitly when the model chooses between a clinical token and a time token [5, 6]. FERMAT differs from both in how it treats events on the same date. Delphi-2M derives the waiting time from the event logits; FERMAT instead uses a separate head that predicts whether the next event falls on the current date and, for an event on a later date, a separate time head that models the waiting time in continuous time rather than in predefined intervals. For record prediction, generation uses the same-day and different-day attention masks applied in training. Our main contribution is empirical: because one pretrained model supplies both applications, the comparison holds the model, patients, observed histories, and outcome definitions fixed, and both applications are evaluated with the same adjustment for incomplete follow-up.

## 2 Study design and methods

### 2.1 FERMAT pretraining and evaluation cohorts

FERMAT was pretrained on longitudinal electronic health records represented in the Observational Medical Outcomes Partnership (OMOP) Common Data Model. The source hospital dataset covered October 2004 to December 2023. The corpus contained 3,773,556 patients and 1,238,329,877 tokenized events, divided by patient into training (2,642,237), validation (565,534), and test (565,785) splits.

Each patient’s records were ordered by date and represented as clinical tokens with an event type and age in days. Diagnoses, prescriptions, procedures, and discretized laboratory measurements provided clinical context. The model combined token and event-type embeddings with a continuous age encoding, allowing multiple items to share the same recorded date. FERMAT is a 65.22-million-parameter autoregressive transformer with a context of up to 512 tokens. Its event output predicts clinical items, while separate time heads estimate whether the next event occurs on the same day and the waiting time to an event on a later date. Appendix A specifies the input domains, prediction targets, and training objective.

Training comprised 100,000 event-prediction steps followed by 100,000 steps with an additional time-prediction loss weighted by 0.3. The two-stage schedule and the weight of 0.3 were selected in preliminary diagnostic runs on 1% of the training data, in which the time loss was added to a model first trained on the event loss alone. The checkpoint with the best validation objective, at step 93,000 of the second stage, was used in both applications. The final observed hidden state supplied the patient representation for task-specific predictors, with FERMAT held fixed. Generated trajectories were obtained by repeatedly predicting an event and its timing and extending the observed history with the generated record.

Eligible index dates had a recorded diagnosis, prescription, procedure, or laboratory test and fell at least two calendar years after the patient’s first clinical record. For each patient, we used the first eligible date in 2018 and the last 512 tokens available through that date. Disease-risk predictors used the eligible training and validation pools, and test patients were randomly sampled from the held-out test pool. Record prediction used random subsets of the training and validation pools and a subset of the disease-risk test sample selected by its random order. Within each evaluation, both applications used the same test patients, history cutoff, and outcome definitions. Sampling details appear in Appendix A.

### 2.2 Estimating disease risk over time

We selected 31 diseases with sufficient sample sizes for disease-specific analysis and evaluated incident disease prediction at annual horizons from one to five years in 10,000 test patients. For each disease, patients with a matching diagnosis through the index date were excluded; an incident event was the first subsequent diagnosis matching the disease-specific concept list. The same concept lists identified disease events in observed and generated records (Appendix E).

For annual discrimination, Cox proportional hazards regression models were fitted using age, sex, and log-transformed counts of prior clinical rows and active clinical days, with or without the FERMAT patient representation [9]. Models were selected by validation performance and refitted on training plus validation data. For patient *i* and disease *d*, the representation-based score was 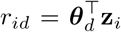 where **z**_*i*_ contains the preprocessed clinical variables and representation and ***θ***_*d*_ contains the fitted coefficients. Each model was fitted to follow-up through five years and supplied the same log-relative hazard score at all annual evaluation horizons.

Generation-based risk was the fraction of usable trajectories containing the disease by horizon *t*:

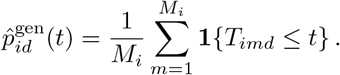

Here, *M*_*i*_ is the number of usable trajectories, and *T*_*imd*_ is the time in years to the first qualifying diagnosis in trajectory *m*, or infinity if none occurs. Fifty trajectories were generated per patient; paths truncated by the token limit before completing follow-up were excluded (Appendix A).

Chronic kidney disease, fatty liver, and diabetes illustrate the detailed probability comparisons. For these examples, weighted logistic models estimated five-year probabilities from the same four clinical variables, with or without the patient representation:

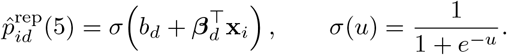

The input **x**_*i*_ contains the clinical variables and representation after training-set preprocessing; *b*_*d*_ and ***β***_*d*_ are fitted coefficients. The clinical-variable baseline omits the representation. Both predictor types used the same fixed regularization across the three diseases (Appendix A). The generated trajectories also supplied annual cohort-mean risks for comparison with observed incidence.

The area under the receiver operating characteristic curve (AUROC) measured discrimination, and the Brier score measured mean squared probability error. Disease before death was a case; death before disease was an observed competing-event noncase. Follow-up reaching the horizon without disease established a noncase; otherwise the outcome was unobserved. Observed outcomes were weighted by the inverse estimated probability of outcome observation, with weights clipped at their first and 99th percentiles. Observation models used the four clinical variables and prior disease indicators. This adjustment assumed conditional independence of outcome observability and the outcome given those variables, an adequate observation model, and positive observation probabilities. For the three detailed disease examples, the effective sample size under the clipped weights was about 93% of the number of patients with an observed outcome (Appendix B), so no small group of patients dominated the weighted estimates. Section 4 discusses how violations could affect the comparisons.

The annual discrimination analysis used prior indicators for the other evaluated diseases in its observation models and counted generated diagnoses through generated death. The detailed probability and incidence analyses used indicators for the other selected disease examples and the first generated diagnosis without an additional generated-death restriction. Figure 1D–F follows the detailed probability-comparison specification. Appendix D gives the complete rules.

**Figure 1.**
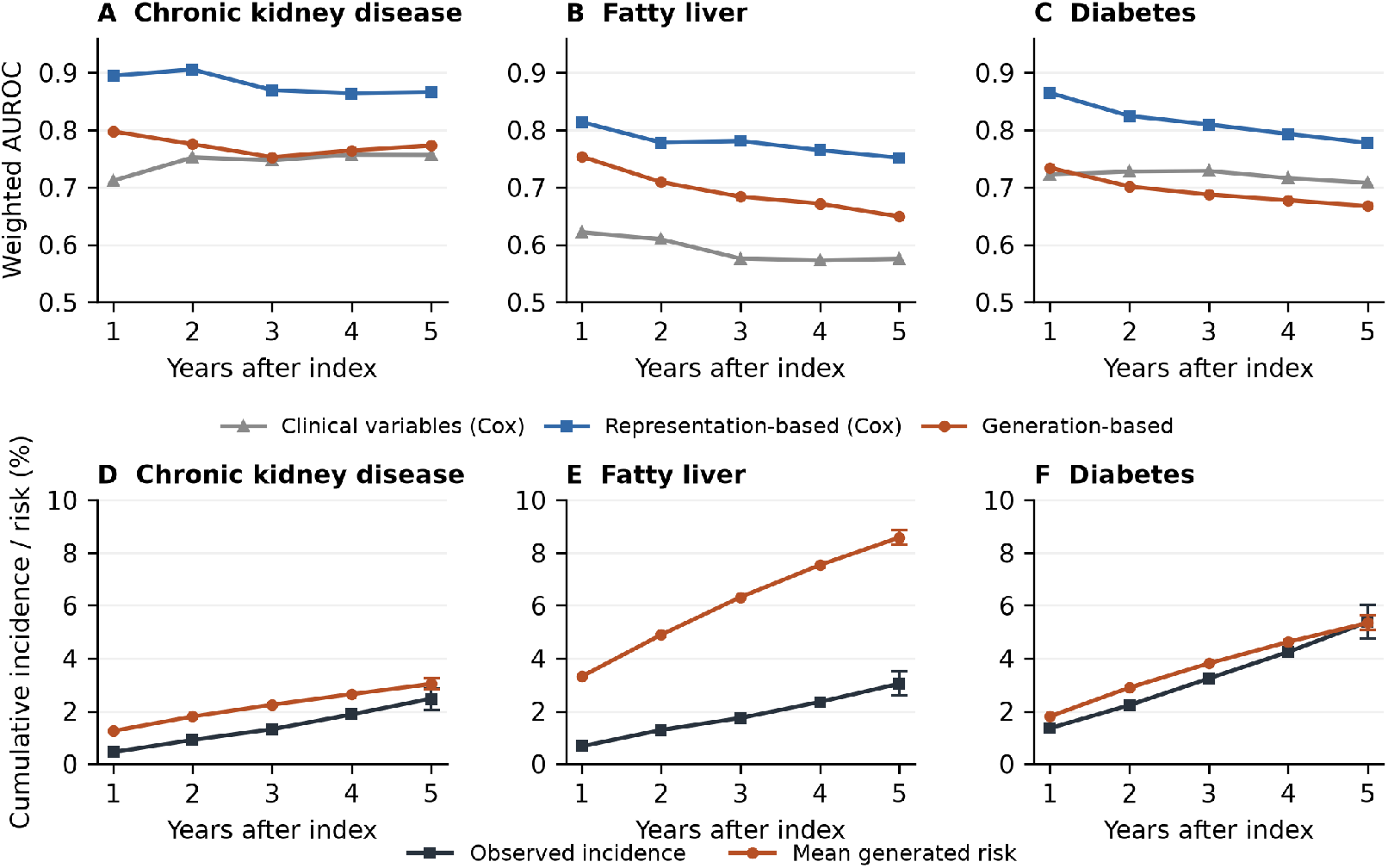
Disease prediction from one to five years. A–C: observation-weighted AUROC point estimates for Cox models using clinical variables alone or clinical variables plus FERMAT representations, compared with generation-based risk estimates. D–F: observation-weighted incidence and cohort-mean risk estimated from generated trajectories for the three selected diseases, with 95% bootstrap intervals at year five. All panels use disease-specific eligible subsets of the 10,000-patient test cohort. Appendix D specifies the observation weights and event-handling rules.

For each disease, the weighted Brier score at horizon *t* was:

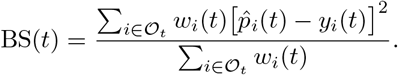

Here, *O*_*t*_ contains disease-eligible patients with an observed outcome at horizon 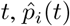 is the predicted probability, *y*_*i*_(*t*) is the binary disease outcome, and *w*_*i*_(*t*) is the clipped inverse observation probability.

### 2.3 Predicting diagnoses, prescriptions, and procedures together

For each patient, we located the first diagnosis, prescription, or procedure recorded on a date after the observed-history cutoff. All diagnosis, prescription, and procedure tokens recorded on that date were targets, including those from different visits. Repeated occurrences of the same token counted once, and their order within the day was ignored. Eligible events had to precede recorded death and occur within the five-year follow-up window. In the training corpus, 80.58% of clinical record days contained multiple distinct diagnosis, prescription, or procedure tokens after within-day deduplication. Patients alive at baseline were included if a target was observed within this window.

For representation-based prediction, linear and one-hidden-layer multilabel heads were fitted; a linear head was selected based on validation Recall@10 (Appendix A). Generation-based prediction used 50 trajectories per patient. For candidate item *c*, the respective ranking scores were:

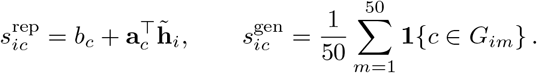

The patient representation 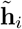 was standardized using training-set means and standard deviations; *b*_*c*_ and **a**_*c*_ are the selected linear head’s fitted parameters. Its score is a logit. The set *G*_*im*_ contains distinct diagnosis, prescription, and procedure tokens on the first clinical date in generated trajectory *m*. An empty set contributes zero for every item and remains in the denominator. Record-prediction inference used separate same-day and different-day event distributions, the full allowed vocabulary, and a temperature of 1.0 (Appendix A).

Each method returned the K highest-scoring distinct items, with K = 1, 2, 5, and 10 and outcome-independent ordering of tied scores. The fitted head and ranking were held fixed across K. For observed set *Y*_*i*_ and predicted set *P*_*i,K*_:

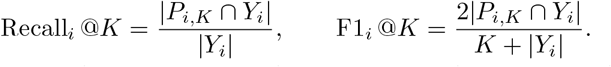

Both metrics were averaged over patients, with Recall@10 as the primary endpoint. Evaluation focused on the clinical items recorded together on the target date (Appendix A).

### 2.4 Statistical analysis

Probability contrasts for the three selected diseases used 2,000 paired bootstrap replicates that resampled patients and then usable paths within patients and refitted the observation model. Risk predictors remained fixed. Record-prediction contrasts used 5,000 paired patient bootstrap replicates, conditioning on fitted heads and generated paths. Annual discrimination comparisons using the Cox models were descriptive point estimates without paired uncertainty intervals; no multiplicity adjustment was applied.

## 3 Results

### 3.1 Patient representations improved disease-risk prediction

Risk scores from representation-augmented Cox models had higher AUROC point estimates than risk estimates from generated trajectories for 30 of 31 diseases at one year and all 31 at each of years 2–5. The one-year exception was ischemic stroke. Figure 1A–C shows annual discrimination for chronic kidney disease, fatty liver, and diabetes; Appendix D reports disease-specific estimates and event counts.

Mean risk estimated from generated trajectories exceeded observed weighted incidence at all annual horizons for fatty liver and chronic kidney disease (Figure 1D–E). For fatty liver, the gap widened from 3.31% versus 0.67% at one year to 8.58% versus 3.04% at five years. Diabetes reached near agreement at five years (5.35% versus 5.38%), despite the lower patient-level AUROC of risk estimates from generated trajectories.

For the three selected disease examples, adding FERMAT representations to the logistic predictor increased AUROC from 0.763 to 0.871 for chronic kidney disease, 0.591 to 0.752 for fatty liver, and 0.715 to 0.784 for diabetes (Figure 2). The representation-based predictors also exceeded generation-based estimates by 0.098–0.115 AUROC, with paired 95% intervals excluding zero (Appendix B).

**Figure 2.**
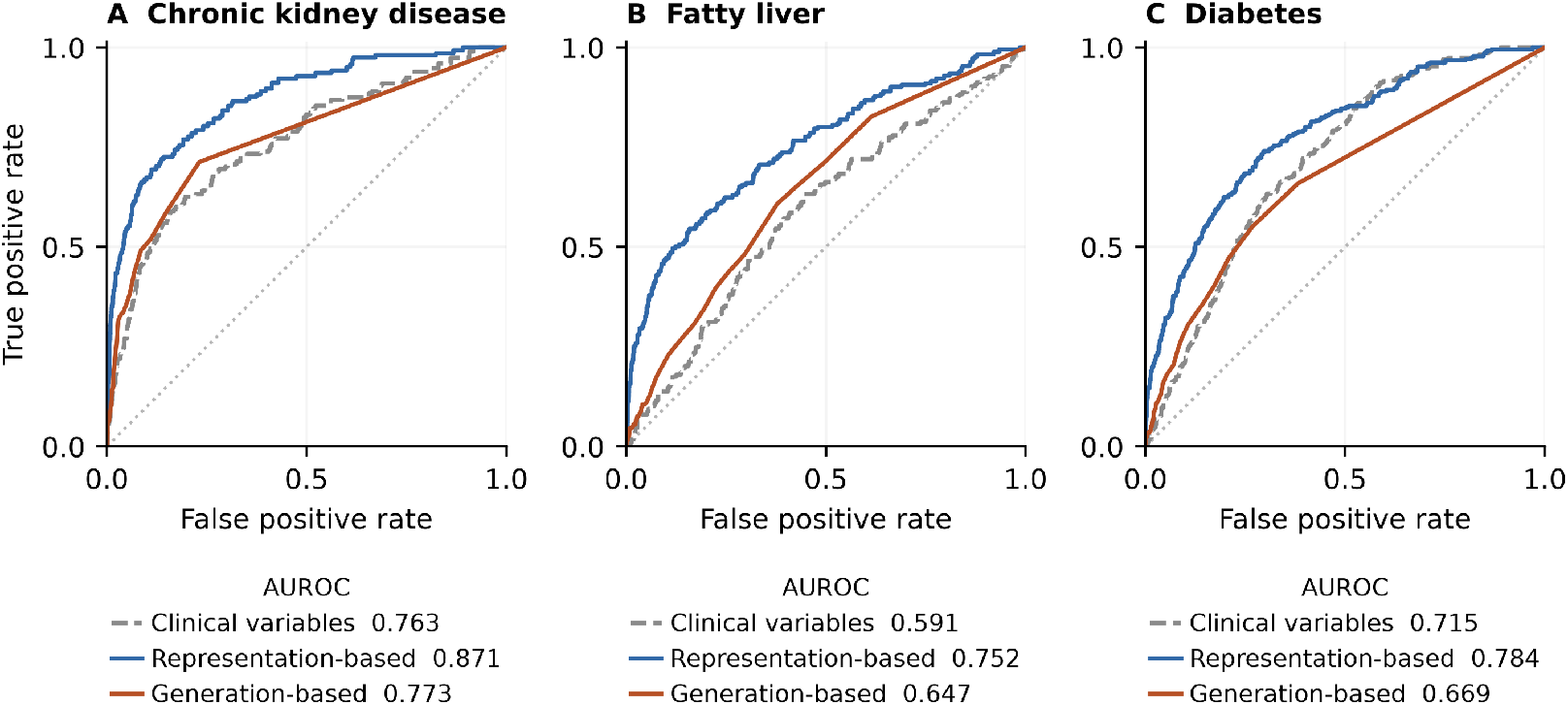
Discrimination of five-year disease risk. Each panel overlays observation-weighted receiver operating characteristic (ROC) curves for three prediction methods on the same disease-specific test patients. The clinical-variable baseline is a logistic predictor using age, sex, and two utilization measures; representation-based prediction adds FERMAT patient representations to those variables. Generation-based prediction uses the fraction of generated trajectories containing the disease within five years. Legends report AUROC. Eligible N was 9,741 for chronic kidney disease, 9,632 for fatty liver, and 8,858 for diabetes; observed outcomes were available for 5,665, 5,578, and 5,082 patients, including 155, 168, and 280 cases, respectively. The dotted diagonal indicates chance discrimination. Paired AUROC differences and their 95% confidence intervals are reported in Appendix B.

Representation-based prediction also had the lowest weighted Brier scores: 0.0204, 0.0273, and 0.0452 for chronic kidney disease, fatty liver, and diabetes, respectively. Corresponding scores were 0.0232, 0.0294, and 0.0496 for the clinical-variable baseline and 0.0249, 0.0468, and 0.0555 for generation-based prediction.

### 3.2 Generation improved prediction of diagnoses, prescriptions, and procedures recorded together

Of 19,992 training, 2,000 validation, and 1,999 test patients alive at baseline, 18,474, 1,867, and 1,862, respectively, had an observed target and were included in the analysis. In the test cohort, the median number of target items was three.

On these 1,862 test patients, generation-based prediction using 50 trajectories per patient achieved a patient-averaged Recall@10 of 71.00%, compared with 55.68% for representation-based prediction (Table 1). The paired improvement was 15.31 percentage points (95% CI, 13.92–16.79); F1@10 improved by 9.73 percentage points (95% CI, 8.92–10.55).

**Table 1.** Prediction of diagnoses, prescriptions, and procedures recorded together. Each method returned its ten highest-scoring clinical items per patient. Values are patient averages in percent.

| Method | Recall@10 | F1@10 |
| --- | --- | --- |
| Representation-based prediction | 55.68 | 28.49 |
| Generation-based prediction (50 trajectories) | **71.00** | **38.22** |

Paired 95% confidence intervals for the difference between generation-based and representation-based prediction included zero at K = 1 and were above zero at K = 2, 5, and 10 for both metrics (Figure 3).

**Figure 3.**
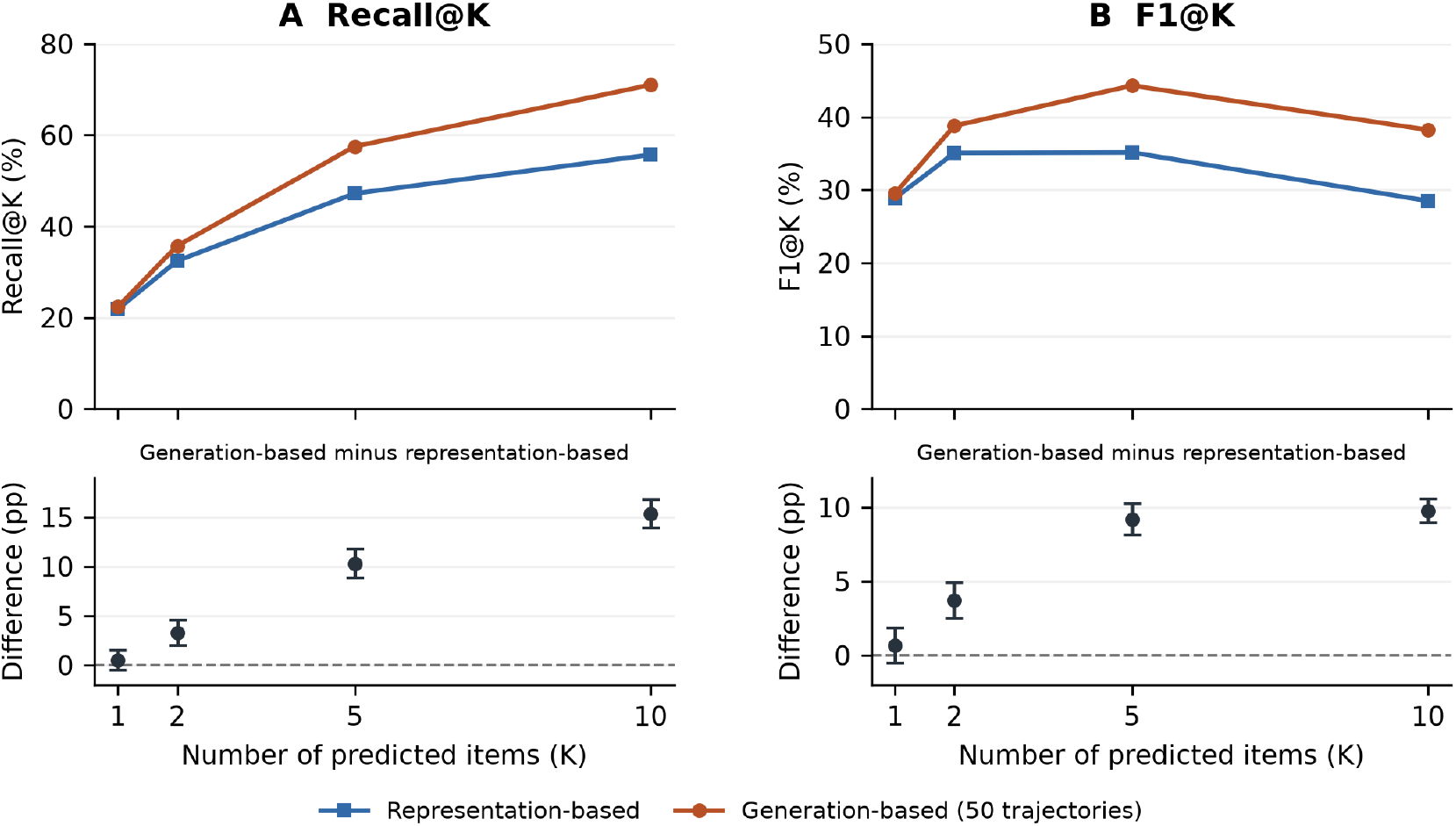
Performance by the number of predicted clinical items. Representation-based prediction and generation-based prediction using 50 generated trajectories are compared on the same 1,862 patients. The horizontal axis gives K, the number of highest-scoring distinct diagnosis, prescription, or procedure items returned for each patient. A: Recall@K, the fraction of items from the actual next recorded day included among those K predictions. B: F1@K, the harmonic mean of precision and recall for the same predictions. Metrics are calculated per patient and then averaged. Within each panel, the upper plot shows mean performance and the lower plot shows the generation-based minus representation-based difference in percentage points. Error bars are 95% percentile confidence intervals from 5,000 paired patient bootstrap replicates, conditional on fitted predictors and generated trajectories; the dashed line marks no difference. Lines connect evaluated K values. Numerical contrasts appear in Appendix C.

## 4 Discussion

FERMAT’s representation-based and generation-based predictions differed in their relative performance across the evaluated tasks. Representation-based prediction performed better in the disease-risk comparisons, whereas generation-based prediction performed better in predicting diagnoses, prescriptions, and procedures recorded together.

The disease-risk findings are consistent with evidence from CLMBR and MOTOR that pretrained patient representations can improve outcome-specific prediction [2, 3]. Adding FERMAT’s patient representations to clinical variables improved prediction, suggesting that pretraining captured disease-relevant information beyond those variables. Predictors incorporating these representations also achieved better discrimination than risk estimates derived from trajectories generated by the same foundation model, with lower probability error in the detailed comparisons.

Generation-based prediction, in turn, achieved significantly better predictive performance than representation-based prediction for diagnoses, prescriptions, and procedures recorded on the first clinical date after the observed history. This extends the findings of Waxler et al., who compared generated encounter contents with predictions that carried forward previously recorded events [5]. Hospital EHRs commonly contain multiple events recorded within a visit or on the same date [8, 11]; same-day events accounted for 84.38% of clinical target tokens in our training data. FERMAT accommodates this structure by determining whether the next event occurs on the same day and generating the event conditional on that determination. As generation proceeds, previously generated events provide additional context for subsequent predictions. We hypothesize that this use of generated context enabled learned associations among diagnoses, prescriptions, and procedures to contribute to the stronger predictive performance observed in this task.

Generation also costs more. A patient representation requires one forward pass over the observed history, whereas each trajectory requires sequential model evaluations; generating the 500,000 disease-risk trajectories (10,000 patients × 50) took about 87 hours on one RTX 5090. With 50 trajectories, a generated risk changes in steps of 0.02, and in record prediction Recall@10 rose from 65.56% with 10 trajectories to 71.00% with 50 (Appendix C). For the risk of a prespecified outcome, a direct predictor was therefore both cheaper and more accurate in our comparisons; generation is better suited to outputs that require a set of co-occurring items or an ordered sequence of events.

Taken together, these findings suggest that the utility of representation-based and generation-based methods varies with the clinical task. The two can be combined in at least three ways, none of which we evaluated. First, a system can assign each request to the better application: representation-based predictors for the risk of a prespecified outcome and generation for next-date item sets or event sequences. Second, summaries of generated trajectories, such as the generated risk or the number of generated events, can be added as inputs to a representation-based predictor, so that the fitted predictor determines how much weight generation receives. Third, representation-based risk estimates can be used to recalibrate generated trajectories, for example by reweighting trajectories so that their cumulative incidence matches the representation-based estimate; this would retain the generated event sequences while correcting overestimation such as that observed for fatty liver and chronic kidney disease. Future work could also investigate whether these representations and trajectories can support counterfactual learning under alternative treatment scenarios, extending the applications of clinical foundation models beyond the prediction tasks evaluated here.

Several limitations of this study should be considered when interpreting the findings. First, the use of data from a single hospital may limit generalizability to populations and healthcare settings with different clinical and documentation practices; evaluation in other institutions and in linked claims data is needed to test whether the task-dependent pattern holds. Second, the available records may not fully capture patients’ clinical histories. For example, the date of the first recorded diagnosis may differ from the actual date of disease onset or initial diagnosis, and some clinical events may be missing, particularly those occurring outside the source hospital. Third, outcomes were defined by a single first recorded diagnosis, without repeated diagnoses, confirming prescriptions or procedures, or registry linkage (Appendix E). Billing practices can inflate recorded diagnoses. Because the same definitions identified events in observed and generated records, the comparison between applications is internally consistent, but absolute performance refers to recorded diagnoses rather than verified disease. Registry data, rules requiring repeated diagnoses or disease-specific treatment, and phenotyping algorithms would give more reliable case definitions.

Incomplete follow-up further limits outcome ascertainment. Observation weighting assumes that, given age, sex, utilization, and prior disease indicators, whether a patient’s outcome is observed is unrelated to the outcome. This assumption fails if patients who develop disease, or who remain healthy, are more likely to leave the source hospital for reasons not captured by these variables. Diagnoses made at other institutions are missed regardless, which weighting cannot correct. Both applications were evaluated on the same patients with the same weights, so a violation affects both; the effects need not be equal, however, because representation-based scores may encode utilization patterns that also predict continued follow-up. The effective sample sizes indicate that the clipped weights were not extreme, which supports positivity but does not test conditional independence. Fourth, we compared two applications of a fixed pretrained model and did not evaluate supervised fine-tuning, for example with low-rank adaptation, which updates the model’s parameters for a target task. Fine-tuning is a natural third application of a foundation model and could change the relative performance in either task. Nevertheless, FERMAT demonstrates how a single clinical foundation model can support prediction through representations and generated trajectories with task-dependent strengths, motivating further work to combine these approaches and extend them to broader clinical applications.

## 5 Conclusion

FERMAT supports clinical prediction through representation-based and generation-based applications of the same pretrained model. Representation-based prediction performed better in the disease-risk comparisons, whereas generation-based prediction performed better in predicting diagnoses, prescriptions, and procedures recorded together. These findings demonstrate task-dependent strengths within a single clinical foundation model and provide a basis for exploring how its representations and generated trajectories can be used and combined to address different clinical prediction needs.

### Research ethics and data availability

This study was conducted after obtaining a review exemption from the Institutional Review Board of Seoul National University Hospital (IRB No. 2608-043-1768). Patient-level data remain restricted.

## Data Availability

All data used in the present study are available upon reasonable request to the authors, subject to Institutional Review Board approval and institutional data access procedures.

## A Implementation and reporting details

The transformer contains ten layers, each with ten attention heads and a 640-dimensional hidden state. Its vocabulary comprises 24,435 tokens, and its maximum context length is 512 tokens.

Input tokens represent clinical concepts and discretized measurements. Context includes diagnoses, prescriptions, procedures, laboratory measurements, sex, death, and available genomic annotations. Numerical laboratory values were discretized using training-data quantiles. Token and event-type embeddings are added to a sinusoidal encoding of age in days. The supervised event targets comprise diagnoses, prescriptions, procedures, and death; laboratory and metadata tokens contribute context. Event prediction uses causal attention with additional masking of events sharing the target date.

Event prediction uses cross-entropy. The time objective combines binary cross-entropy for whether the next event is on the same date with exponential waiting-time negative log-likelihood for events on later dates. The same-day head uses a causal representation independent of the target date; a separate linear time head predicts the waiting-time rate. In the second training stage, the joint objective is 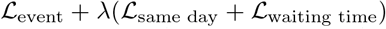 with *λ* increasing to 0.3 over the first 200 steps. Event and time losses use their respective eligible targets.

For disease-risk evaluation, patients in the development sample were excluded from the test sampling pool. Eligible patients were placed in random order, and the first 10,000 formed the test cohort. The record-prediction test sample used the first 2,000 patients in that order. Its training and validation samples were drawn without replacement from the corresponding eligible pools, with sample sizes of 20,000 and 2,000 before baseline-death and target-availability exclusions. Disease-risk predictor training used the eligible training pool, and validation used the eligible validation pool.

Foundation-model training used a learning rate of 0.0002, a minimum learning rate of 0.00002, dropout of 0.1, and weight decay of 0.1. Training used a microbatch of eight, gradient accumulation of 16, BF16 precision, and an NVIDIA RTX 5090. Pretraining took about 49 hours of wall-clock time on this single GPU (about 49 GPU-hours).

The disease-risk logistic predictors were fitted with weighted binary cross-entropy using AdamW for 200 steps at a learning rate of 0.03. Regularization was fixed at C = 10, corresponding to weight decay 0.00001 under the mapping 0.0001/C, for both predictor types across all three diseases. Training-set medians, means, and standard deviations defined imputation and scaling.

The prediction vocabulary comprises 17,460 diagnosis, prescription, and procedure tokens. The event-set head uses unweighted multilabel binary cross-entropy, AdamW with a learning rate of 0.001 and weight decay of 0.01, a batch size of 256, and at most 60 epochs. The multilayer perceptron (MLP) has one hidden layer with 256 units. Training stops after ten epochs without improvement in validation Recall@10; validation binary cross-entropy breaks ties. Linear and MLP heads were each trained from three random initializations. One linear head was used for the comparisons in the main text. Test Recall@10 across the three linear heads is 55.76%, 55.68%, and 55.98%; across the three MLP heads it is 52.17%, 51.77%, and 52.28%.

The disease-risk sampler uses top-50 sampling, temperature 0.8, a same-day repetition penalty that subtracts 1.0 from the logits of tokens already present on that day, and a 2,048-new-token limit. After excluding trajectories truncated before completing follow-up, one test patient had 49 usable trajectories and each remaining patient had 50. The record-prediction sampler uses the full allowed clinical vocabulary, temperature 1.0, no same-day repeat penalty, and independent path-specific random streams. Generation stopped upon reaching death, the horizon, or the token limit. With 50 paths, expected Recall@10 under equally likely score-tie orders is 70.96%, close to the fixed-tie result of 71.00%.

Record-prediction inference computes three hidden states from the same observed prefix: a plain causal state and two states with the training-compatible same-day and different-day attention masks. At the last observed position, the branch masks use the last observed age and that age plus one day, respectively; only equality versus inequality is needed to select the mask. A sigmoid same-day head applied to the plain state supplies the mixture weight. The shared event output layer supplies a softmax distribution from each branch state after masking disallowed outputs. Sequential generation samples a branch, an event from that branch, and a waiting time from the model, then repeats on the extended prefix.

For record-prediction evaluation, generated elapsed days were mapped to floor(offset + 1) before extracting sets, so raw day-zero events contributed to the first future day.

Disease outcomes were ascertained from the source diagnosis records using the definitions in Appendix E.

The table reports disease-specific training and validation cohort sizes. Each cell reports eligible patients followed by patients with an observed outcome in that split.

| Disease | Training eligible / observed | Validation eligible / observed |
| --- | --- | --- |
| Chronic kidney disease | 256,536 / 148,048 | 54,858 / 31,925 |
| Fatty liver | 253,697 / 145,782 | 54,319 / 31,461 |
| Diabetes | 235,670 / 133,947 | 50,477 / 28,974 |

## B Disease-risk uncertainty and follow-up

The paired AUROC differences below were calculated by subtracting the AUROC of generation-based risk estimates from that of representation-based logistic predictors. The confidence intervals condition on fitted risk predictors and account for patient and within-patient path resampling.

| Disease | AUROC difference | Paired 95% CI |
| --- | --- | --- |
| Chronic kidney disease | +0.098 | +0.079 to +0.152 |
| Fatty liver | +0.105 | +0.063 to +0.158 |
| Diabetes | +0.115 | +0.094 to +0.159 |

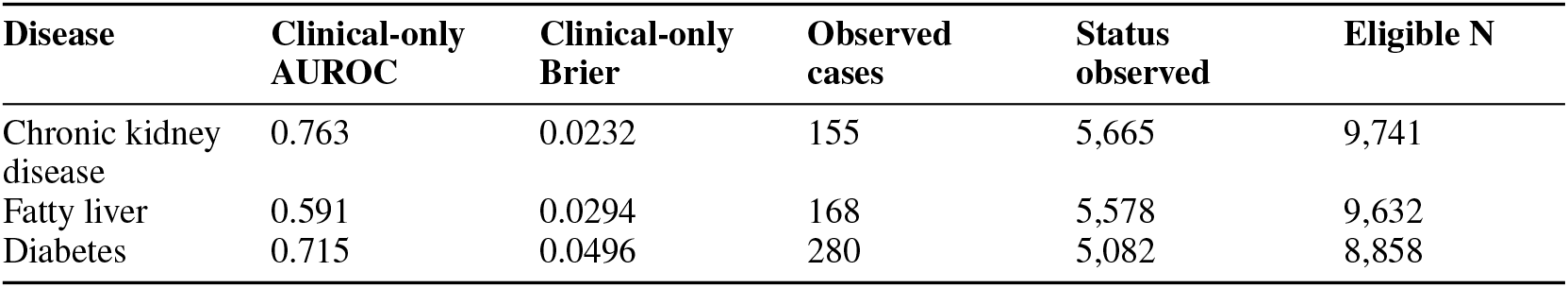

Clinical-only denotes the age, sex, and utilization logistic predictor. Deaths before disease numbered 60, 75, and 50, respectively. The corresponding effective sample sizes under observation weights were 5,264, 5,213, and 4,705. Outcome status at five years was unobserved for the remaining eligible patients.

| Disease | Year | Observation-weighted incidence (%) | Mean estimated risk (%) |
| --- | --- | --- | --- |
| Chronic kidney disease | 1 | 0.45 | 1.25 |
| Chronic kidney disease | 2 | 0.91 | 1.80 |
| Chronic kidney disease | 3 | 1.32 | 2.25 |
| Chronic kidney disease | 4 | 1.88 | 2.65 |
| Chronic kidney disease | 5 | 2.47 | 3.03 |
| Fatty liver | 1 | 0.67 | 3.31 |
| Fatty liver | 2 | 1.29 | 4.89 |
| Fatty liver | 3 | 1.74 | 6.32 |
| Fatty liver | 4 | 2.35 | 7.54 |
| Fatty liver | 5 | 3.04 | 8.58 |
| Diabetes | 1 | 1.36 | 1.81 |
| Diabetes | 2 | 2.23 | 2.90 |
| Diabetes | 3 | 3.24 | 3.81 |
| Diabetes | 4 | 4.25 | 4.63 |
| Diabetes | 5 | 5.38 | 5.35 |

The table compares mean risk estimated from generated trajectories with observation-weighted cohort incidence.

## C Supplementary record-prediction analyses

### C.1 Supplementary comparisons

Training-population frequency and reuse of the last observed record provided simple reference rankings. The reuse control ranked items from the last recorded day by training frequency and filled remaining positions from the rest of the clinical vocabulary. These methods achieved Recall@10 of 8.73% and 46.31%, respectively.

Aggregation of 10, 20, and 50 trajectories achieved Recall@10 of 65.56%, 68.85%, and 71.00%, respectively. The difference between 50 and 10 trajectories was 5.43 percentage points (95% CI, 4.55–6.32).

### C.2 Metrics and paired comparisons

K denotes the number of highest-scoring clinical items selected per patient. The tables compare representation-based prediction and generation-based prediction with different numbers of generated trajectories.

**Recall@K (%)**.

| Method | K=1 | K=2 | K=5 | K=10 |
| --- | --- | --- | --- | --- |
| Representation-based | 21.89 | 32.43 | 47.21 | 55.68 |
| Generation-based (10 paths) | 20.79 | 34.31 | 54.97 | 65.56 |
| Generation-based (20 paths) | 21.72 | 35.00 | 56.69 | 68.85 |
| Generation-based (50 paths) | 22.38 | 35.72 | 57.51 | 71.00 |

**F1@K (%)**.

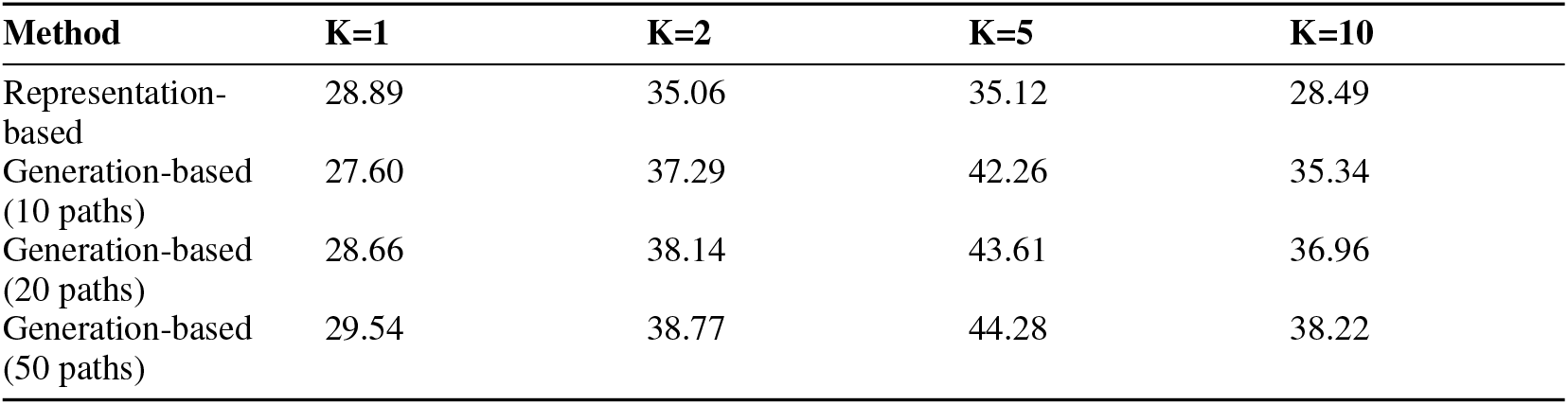

**Generation-based (50 paths) minus representation-based Recall@K: percentage-point difference [95% CI]**.

| K | Difference [95% CI] |
| --- | --- |
| 1 | +0.49 [-0.52, +1.52] |
| 2 | +3.29 [+1.98, +4.57] |
| 5 | +10.30 [+8.85, +11.76] |
| 10 | +15.31 [+13.92, +16.79] |

**Generation-based (50 paths) minus representation-based F1@K: percentage-point difference [95% CI]**.

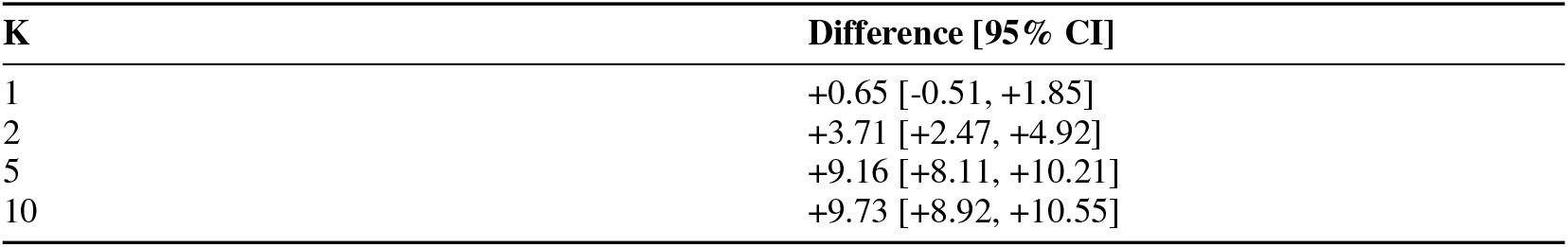

**Precision@K (%)**.

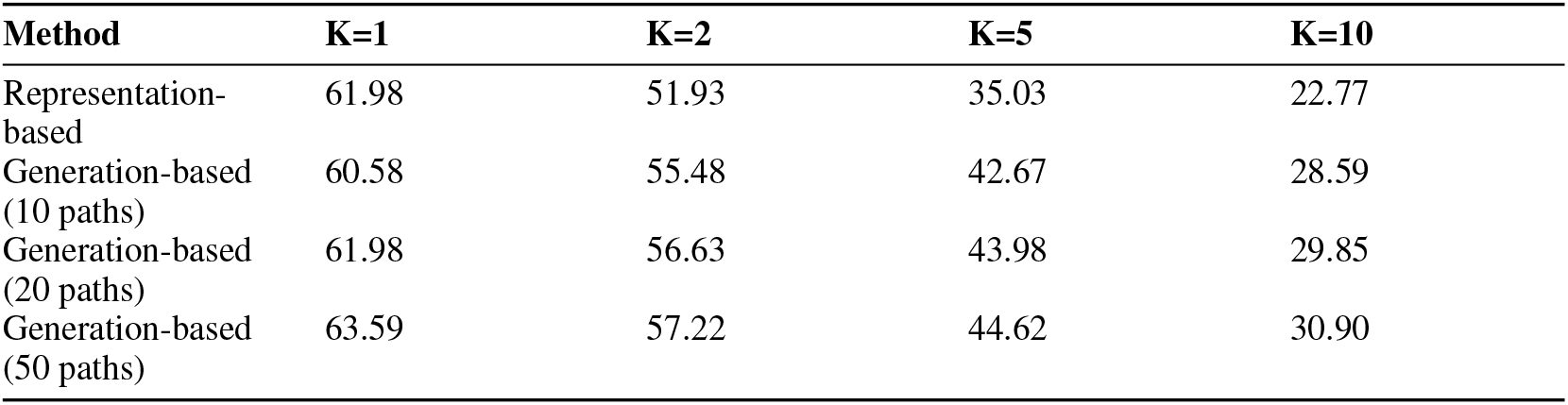

**Hit@K (%)**.

| Method | K=1 | K=2 | K=5 | K=10 |
| --- | --- | --- | --- | --- |
| Representation-based | 61.98 | 69.66 | 78.41 | 82.38 |
| Generation-based (10 paths) | 60.58 | 69.50 | 79.11 | 83.03 |
| Generation-based (20 paths) | 61.98 | 70.46 | 80.50 | 85.77 |
| Generation-based (50 paths) | 63.59 | 70.84 | 80.77 | 86.47 |

Precision@K is the patient-averaged fraction of the K predicted items that are correct. Hit@K is the fraction of patients with at least one correct prediction.

## D Disease discrimination across annual horizons

Discrimination for 31 diseases was evaluated at 12, 24, 36, 48, and 60 calendar months in the 10,000-patient test cohort. Figure S1 shows all point-estimate contrasts with risk estimates from generated trajectories; Table S1 gives eligible populations and annual event counts. Comparisons are correlated across diseases and years. Seven one-year outcomes had fewer than 20 observed cases, and chronic hepatitis B had fewer than 20 at every annual horizon; these cells are explicitly marked and no disease-wise significance is inferred.

The Cox proportional hazards models [9] were fitted by maximizing the L2-penalized partial likelihood, using the Breslow approximation for tied event times. Patients were followed until a qualifying diagnosis or the earliest of death, the last clinical record, and five years after index; death without disease was censored when fitting the Cox models. Penalties were selected by five-year validation observation-weighted AUROC, with smaller penalties breaking ties: {0, 0.0001, 0.001} for clinical variables, and {0.0001, 0.001, 0.01} with representations. Preprocessing and coefficients were then refitted on training plus validation patients. The linear predictor from each fitted Cox model was used as the risk score for discrimination at every annual horizon.

The annual evaluator used each patient’s exact calendar horizon. A recorded disease counted only if it occurred by the horizon, no later than death, and within recorded follow-up. Death before disease and within follow-up counted as an observed noncase; otherwise follow-up had to reach the horizon. Same-day disease and death gave precedence to disease. For annual discrimination, the observation model used L2-regularized logistic regression (C = 1) with the four baseline variables and prior indicators for the other 30 diseases. These predictors were standardized and median-imputed within the eligible analysis population. Inverse observation weights were clipped at their first and 99th percentiles. At each horizon, these observation models estimated the probabilities used to construct the evaluation weights. The observation model included indicators for the other two diseases in the detailed probability comparisons and for the other 30 diseases in the annual discrimination analysis.

Generation used day-plus-one trajectories, retaining days 1–1,826. Paths truncated by the token cap before the horizon were excluded from the denominator. The annual evaluator counted disease events through generated death, including equal-day events. The three-disease analysis used the first generated disease date without an additional generated-death restriction. Figure 1D–F uses the observation weights from the three-disease analysis.

**Figure S1.**
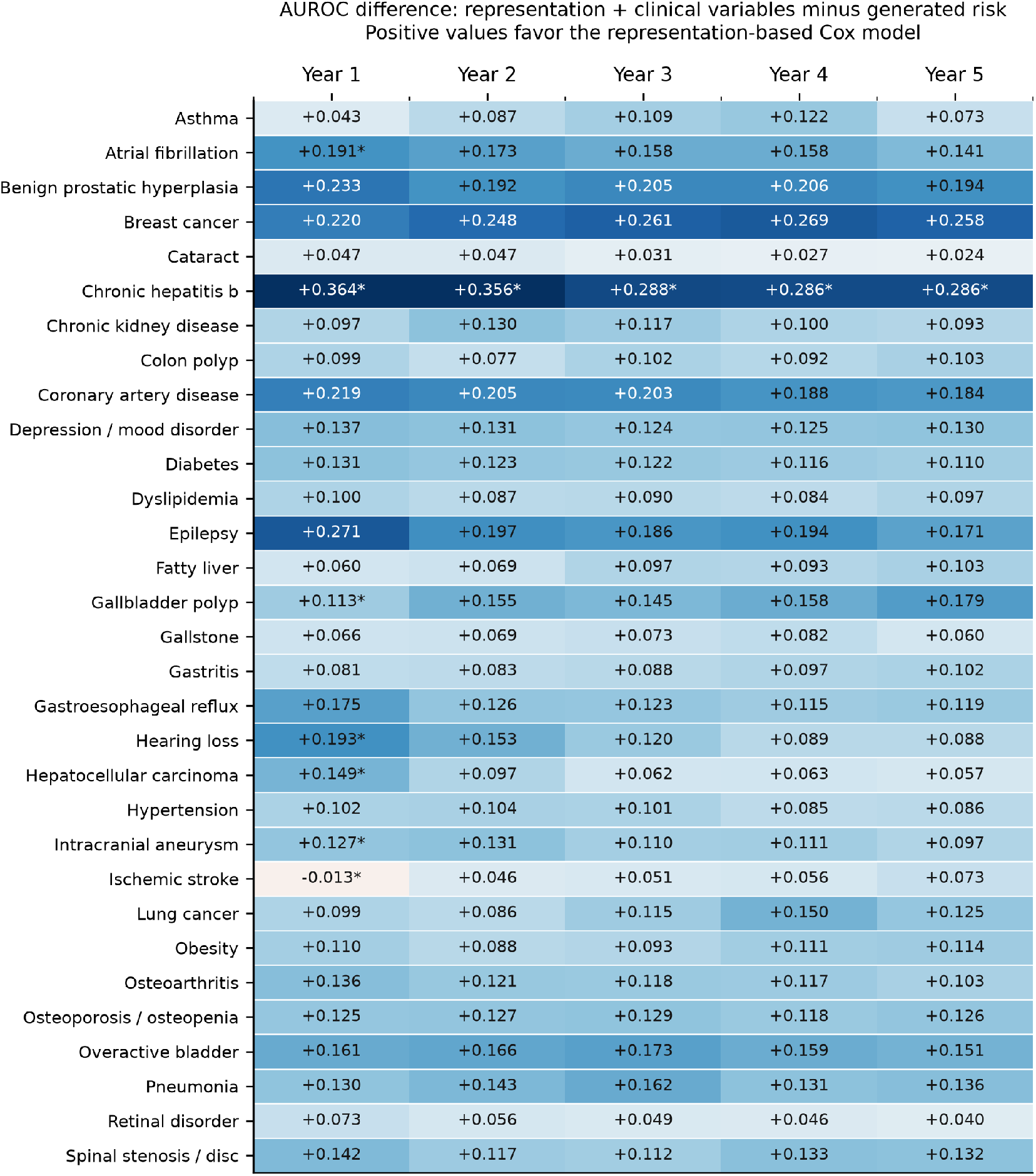
AUROC difference between representation-augmented Cox models and generation-based risk estimates. Positive differences favor representations. Columns are years after the prediction index. An asterisk marks fewer than 20 observed disease cases. These are point estimates, without paired uncertainty intervals or multiplicity adjustment. The one negative cell is ischemic stroke at one year.

**Table S1.** Eligible patients and observed disease cases at annual horizons. Counts refer to patients with a qualifying first recorded diagnosis. Figure S1 marks comparisons with fewer than 20 cases.

| Disease | Eligible N | Y1 cases | Y2 cases | Y3 cases | Y4 cases | Y5 cases |
| --- | --- | --- | --- | --- | --- | --- |
| Asthma | 9,719 | 21 | 33 | 42 | 49 | 57 |
| Atrial fibrillation | 9,800 | 18 | 31 | 42 | 60 | 76 |
| Benign prostatic hyperplasia | 9,475 | 41 | 81 | 100 | 119 | 159 |
| Breast cancer | 9,631 | 22 | 30 | 43 | 54 | 63 |
| Cataract | 9,311 | 62 | 131 | 170 | 224 | 272 |
| Chronic hepatitis B | 9,974 | 3 | 4 | 5 | 5 | 5 |
| Chronic kidney disease | 9,741 | 37 | 72 | 96 | 126 | 155 |
| Colon polyp | 9,629 | 74 | 131 | 173 | 210 | 244 |
| Coronary artery disease | 9,701 | 36 | 83 | 120 | 162 | 190 |
| Depression or mood disorder | 9,381 | 94 | 132 | 166 | 207 | 236 |
| Diabetes | 8,858 | 99 | 152 | 203 | 243 | 280 |
| Dyslipidemia | 8,514 | 150 | 284 | 382 | 467 | 543 |
| Epilepsy | 9,648 | 22 | 27 | 32 | 40 | 46 |
| Fatty liver | 9,632 | 55 | 96 | 120 | 146 | 168 |
| Gallbladder polyp | 9,849 | 16 | 32 | 37 | 56 | 69 |
| Gallstone | 9,787 | 27 | 51 | 64 | 76 | 88 |
| Gastritis | 8,620 | 186 | 288 | 399 | 461 | 495 |
| Gastroesophageal reflux | 9,349 | 107 | 198 | 303 | 385 | 446 |
| Hearing loss | 9,789 | 12 | 21 | 38 | 57 | 77 |
| Hepatocellular carcinoma | 9,817 | 12 | 28 | 40 | 55 | 66 |
| Hypertension | 8,534 | 118 | 204 | 263 | 324 | 385 |
| Intracranial aneurysm | 9,882 | 14 | 26 | 34 | 51 | 65 |
| Ischemic stroke | 9,643 | 19 | 28 | 48 | 67 | 76 |
| Lung cancer | 9,844 | 25 | 30 | 44 | 57 | 73 |
| Obesity | 9,836 | 32 | 59 | 62 | 72 | 84 |
| Osteoarthritis | 9,254 | 73 | 132 | 185 | 227 | 267 |
| Osteoporosis or osteopenia | 9,519 | 80 | 144 | 186 | 260 | 309 |
| Overactive bladder | 9,832 | 30 | 66 | 87 | 104 | 122 |
| Pneumonia | 9,711 | 43 | 79 | 98 | 113 | 143 |
| Retinal disorder | 9,191 | 50 | 114 | 166 | 221 | 270 |
| Spinal stenosis or disc | 9,401 | 65 | 122 | 171 | 215 | 244 |

## E Recorded disease definitions

Outcomes were defined by the first matching diagnosis concept in the source records. In total, 756 phenotype–concept mappings defined the disease outcomes. A matching diagnosis through the index excluded that patient from the corresponding disease analysis; the same lists were used to identify subsequent diagnoses and generated disease tokens. No repeated-claim requirement or additional medication or laboratory confirmation was applied. These outcomes were defined by the first recorded diagnosis, which may not coincide with disease onset.

The following tables list all diagnosis concepts used for the three selected diseases, with their OMOP concept identifiers and names.

### E.1 Chronic kidney disease

| OMOP concept ID | Concept name |
| --- | --- |
| 193782 | End-stage renal disease |
| 197921 | Renal osteodystrophy |
| 198185 | Chronic renal failure |
| 443597 | Chronic kidney disease stage 3 |
| 443601 | Chronic kidney disease stage 2 |
| 443611 | Chronic kidney disease stage 5 |
| 443612 | Chronic kidney disease stage 4 |
| 443614 | Chronic kidney disease stage 1 |
| 443961 | Anemia of chronic renal failure |
| 4030520 | End stage renal failure on dialysis |
| 4128067 | Acute-on-chronic renal failure |
| 43531562 | Chronic kidney disease stage 5 due to type 2 diabetes mellitus |
| 43531578 | Chronic kidney disease due to type 2 diabetes mellitus |
| 45769903 | Chronic kidney disease stage 5 due to type 1 diabetes mellitus |
| 45773688 | Chronic kidney disease due to type 1 diabetes mellitus |
| 46271022 | Chronic kidney disease |

### E.2 Fatty liver

| OMOP concept ID | Concept name |
| --- | --- |
| 193256 | Alcoholic fatty liver |
| 4026131 | Non-alcoholic fatty liver |
| 4059290 | Steatosis of liver |
| 37396531 | Acute fatty liver of pregnancy |
| 40484532 | Nonalcoholic steatohepatitis |

### E.3 Diabetes

| OMOP concept ID | Concept name |
| --- | --- |
| 193323 | Neonatal diabetes mellitus |
| 195771 | Secondary diabetes mellitus |
| 201254 | Type 1 diabetes mellitus |
| 201820 | Diabetes mellitus |
| 201826 | Type 2 diabetes mellitus |
| 443412 | Type 1 diabetes mellitus without complication |
| 4008576 | Diabetes mellitus without complication |
| 4024659 | Gestational diabetes mellitus |
| 4034960 | Secondary endocrine diabetes mellitus |
| 4058243 | Diabetes mellitus during pregnancy, childbirth and the puerperium |
| 4099653 | Steroid-induced diabetes mellitus without complication |
| 4166381 | Maternal diabetes mellitus with hypoglycemia affecting fetus OR newborn |
| 4178452 | Diabetes mellitus associated with pancreatic disease |
| 4193704 | Type 2 diabetes mellitus without complication |
| 4202383 | Drug-induced diabetes mellitus |
| 4230254 | Type 2 diabetes mellitus in nonobese |
| 4304377 | Type 2 diabetes mellitus in obese |
| 4322638 | Diabetes mellitus AND insipidus with optic atrophy AND deafness |
| 4327944 | Malnutrition related diabetes mellitus |
| 40482883 | Posttransplant diabetes mellitus |
| 43531007 | Pre-existing diabetes mellitus in pregnancy |
| 43531008 | Pre-existing type 1 diabetes mellitus in pregnancy |
| 43531010 | Pre-existing type 2 diabetes mellitus in pregnancy |
| 43531640 | Maturity-onset diabetes of the young |
| 44787902 | Lipoatrophic diabetes mellitus without complication |
| 44793113 | Diabetes mellitus with multiple complications |

